# Estimation of secondary household attack rates for emergent SARS-CoV-2 variants detected by genomic surveillance at a community-based testing site in San Francisco

**DOI:** 10.1101/2021.03.01.21252705

**Authors:** James Peng, Sabrina A Mann, Anthea M Mitchell, Jamin Liu, Matthew T. Laurie, Sara Sunshine, Genay Pilarowski, Patrick Ayscue, Amy Kistler, Manu Vanaerschot, Lucy M. Li, Aaron McGeever, Eric D. Chow, IDseq Team, Carina Marquez, Robert Nakamura, Luis Rubio, Gabriel Chamie, Diane Jones, Jon Jacobo, Susana Rojas, Susy Rojas, Valerie Tulier-Laiwa, Douglas Black, Jackie Martinez, Jamie Naso, Joshua Schwab, Maya Petersen, Diane Havlir, Joseph DeRisi

## Abstract

**Background:** Sequencing of the SARS-CoV-2 viral genome from patient samples is an important epidemiological tool for monitoring and responding to the pandemic, including the emergence of new mutations in specific communities.

**Methods:** SARS-CoV-2 genomic sequences were generated from positive samples collected, along with epidemiological metadata, at a walk-up, rapid testing site in the Mission District of San Francisco, California during November 22-December 2, 2020 and January 10-29, 2021. Secondary household attack rates and mean sample viral load were estimated and compared across observed variants.

**Results:** A total of 12,124 tests were performed yielding 1,099 positives. From these, 811 high quality genomes were generated. Certain viral lineages bearing spike mutations, defined in part by L452R, S13I, and W152C, comprised 54.9% of the total sequences from January, compared to 15.7% in November. Household contacts exposed to “West Coast” variants were at higher risk of infection compared to household contacts exposed to lineages lacking these variants (0.357 vs 0.294, RR=1.29; 95% CI:1.01-1.64). The reproductive number was estimated to be modestly higher than other lineages spreading in California during the second half of 2020. Viral loads were similar among persons infected with West Coast versus non-West Coast strains, as was the proportion of individuals with symptoms (60.9% vs 64.1%).

**Conclusions:** The increase in prevalence, relative household attack rates, and reproductive number are consistent with a modest transmissibility increase of the West Coast variants; however, additional laboratory and epidemiological studies are required to better understand differences between these variants.

**Summary:** We observed a growing prevalence and elevated attack rate for “West Coast” SARS-CoV-2 variants in a community testing setting in San Francisco during January 2021, suggesting its modestly higher transmissibility.

## INTRODUCTION

Genomic surveillance during the SARS-CoV-2 pandemic is a critical source of situational intelligence for epidemiological control measures, including outbreak investigations and detection of emergent variants. Countries with robust, unified public health systems and systematic genomic surveillance have been able to rapidly detect SARS-CoV-2 variants with increased transmission characteristics, and mutations that potentially subvert both naturally acquired or vaccination-based immunity (e.g. COVID-19 Genomics UK Consortium). Examples include the rapidly spreading B.1.1.7 lineage documented in the UK and the B.1.351 lineage described from South Africa. The P.1/P.2 lineages originally discovered in Brazil harbor the E484K mutation within the spike protein which, along with other mutations, is associated with reduced neutralization in laboratory experiments[1–4].

In the US, genomic surveillance is sparse relative to the number of confirmed cases (27.8 million as of Feb 20, 2021), with 123,672 genomes deposited in the GISAID database, representing only 0.4% of the total reported cases. Despite the lower rates of overall US genomic surveillance, independent local programs and efforts have contributed to our understanding of variant emergence and spread[5]. The appearance of new nonsynonymous mutations highlight the utility of this approach in the US[6].

Genomic sequencing of SARS-CoV-2 in California has predominantly been conducted by academic researchers and non-profit biomedical research institutions (for example, the Chan Zuckerberg Biohub and Andersen Lab at the Scripps Research Institute) in conjunction with state and local public health partners. These efforts identified an apparent increase in the prevalence of lineages B.1.427 and B.1.429 (“West Coast” variant), which share S gene nonsynonymous mutations at sites 13, 152, 452, and 614, during December 2020 to February 2021 when California was experiencing the largest peak of cases observed during the pandemic. While the cluster of mutations was first observed in a sample from May 2020, these variants rose from representing <1% of the consensus genomes recovered from California samples collected in October 2020 (5/546; 0.91%) to over 50% of those collected during January, 2021 (2,309/4,305; 53.6%; GISAID accessed February 20, 2021).

The majority of efforts in the US utilize samples sent for clinical analysis from symptomatic individuals or outbreaks, introducing selection bias making interpretation of trends, such as the rise in lineage prevalence. Clinical remnant samples are most often delinked from case information, thus eliminating the possibility of evaluating genotypes with detailed geospatial information, household information, and other metadata useful for investigation of transmission dynamics.

Sequencing cases identified during intensive, longitudinal community-based testing may help address both limitations. Here, we describe an investigation of the prevalence of the West Coast variants as well as other variants among persons tested at a community testing site situated in the Mission District neighborhood, San Francisco with high COVID-19 incidence during two periods: November 22-December 2, 2020 and January 10-29, 2021. Using metadata collected at the testing site and Supplementary household testing, we estimated secondary household attack rate with respect to viral genotype to evaluate relative transmissibility of identified variants.

## METHODS

### Study setting and population

From January 10-29, 2021, BinaxNOW™ rapid antigen tests were performed at the 24th & Mission BART (public transit) station in the Mission District of San Francisco, a setting of ongoing community transmission, predominantly among Latinx persons[7]. Tests for SARS-CoV-2 were performed free of charge on a walk-up, no-appointment basis, including persons ≥ 1 year of age, regardless of symptoms through Unidos en Salud, an academic, community (Latino Task Force) and city partnership. Certified lab assistants collected 2 bilateral anterior nasal swabs. The first was tested with BinaxNOW™, immediately followed by a separate bilateral swab for SARS-CoV-2 genomic sequencing[8,9]. Results were reported to participants within 2 hours, and all persons in a household corresponding to a positive BinaxNOW case were offered same-day BinaxNOW testing. All persons testing BinaxNOW positive were offered participation in longitudinal Community Wellness Team support program[10,11].

### SARS-CoV-2 genomic sequence recovery and consensus genome generation

SARS-CoV-2 genomes were recovered using ARTIC Network V3 primers[12] and sequenced on an Illumina NovaSeq platform. Consensus genomes generated from the resulting raw .fastq files using IDseq [13] were used for subsequent analysis. Full details are included in Supplementary Methods.

### Household attack rate analyses

Households (n=327) meeting the following inclusion criteria were eligible for secondary attack rate analyses: 1) ≥1 adult (aged ≥18 years) with a positive BinaxNOW result; 2) ≥1 case in household sequenced; and, 3) ≥2 persons tested with BinaxNOW during the study period.

Households in which sequences represented both West Coast and non-West Coast variants were excluded (n=9). The index was defined as the first adult to test positive. Crude household attack rates, stratified by variant classification, were calculated as i) the proportion of positive BinaxNOW results among tested household contacts; and, ii) as the mean of household-specific secondary attack rate, with 95% CI based on cluster-level bootstrap. Generalized estimating equations were used to fit Poisson regressions, with cluster-robust standard errors and an exchangeable working covariance matrix. Because symptoms and disease severity may be affected by strain, these factors were not included in the a priori adjustment set. We evaluated for overdispersion[14], and conducted sensitivity analyses using targeted maximum likelihood estimation (TMLE) combined with Super Learning to relax parametric model assumptions; influence curve-based standard error estimates used household as the unit of independence[15].

### Bayesian Phylogenetic Analysis

We compared the growth rates of B.1.427 and B.1.429 PANGO lineages against two other lineages, B.1.232 and B.1.243 that had been circulating in California during the latter half of 2020. To do this, we built a Bayesian phylogeny for each lineage in BEAST v.1.10.4 and estimated the effective population size over time using the Bayesian SkyGrid model. We fit an exponential model to the median SkyGrid curve and inferred the reproductive numbers based on the exponential growth rates and generation time estimates from literature. Full analysis details are included in Supplementary Methods.

### Ethics statement

The UCSF Committee on Human Research determined that the study met criteria for public health surveillance. All participants provided informed consent for dual testing.

## RESULTS

### Low-Barrier SARS-CoV-2 Testing and Sequencing

From January 10-29, 8,822 rapid direct antigen tests were performed on 7,696 unique individuals, representing 5,239 households (Supplementary Table 1). Test subjects originated from addresses in 8 Bay Area counties, indicating a wide catchment area (Figure 1). During this time period, there were 885 (10.0%) samples from 863 unique persons that were BinaxNOW positive for SARS-CoV-2 infection. From this set, a total of 165 samples were sequenced for the S gene only, of which 96 had S gene coverage over 92%. In addition, full SARS-CoV-2 genome sequencing was attempted on a total of 674 samples, of which 620 samples resulted in a genome coverage over 92% (Supplementary Table 2, sequences deposited in GISAID). These 716 samples, together with an additional 191 SARS-CoV-2 genome sequences generated from the same testing site during the period of November 22-December 2, 2020[16], had adequate coverage of the full genome or spike protein for further analysis based on S gene sequence (Supplementary Table 3). Classification as either a West Coast variant or a non-West Coast variant was determined for 830 of all samples sequenced.

**Table 1:** Characteristics of households included in the household attack rate analysis, stratified by strain.

|  | Non-West Coast<br>(N=156) | West coast |  |  | Total<br>(N=318) |
| --- | --- | --- | --- | --- | --- |
|  |  | B.1.427<br>(N=78) | B.1.429<br>(N=60) | All West coast<br>(N=162)<sup>+</sup> |  |
| **Race/Ethnicity (most common in household)** |  |  |  |  |  |
| Hispanic/Latinx | 143 (91.7%) | 68 (87.2%) | 58 (96.7%) | 145 (89.5%) | 288 (90.6%) |
| Asian | 5 (3.2%) | 4 (5.1%) | 1 (1.7%) | 8 (4.9%) | 13 (4.1%) |
| White/Caucasian | 4 (2.6%) | 2 (2.6%) | 0 (0%) | 3 (1.9%) | 7 (2.2%) |
| Black or African American | 2 (1.3%) | 2 (2.6%) | 1 (1.7%) | 4 (2.5%) | 6 (1.9%) |
| Other | 2 (1.3%) | 2 (2.6%) | 0 (0%) | 2 (1.2%) | 4 (1.3%) |
| **Has Children** |  |  |  |  |  |
| Does not have children | 105 (67.3%) | 62 (79.5%) | 32 (53.3%) | 109 (67.3%) | 214 (67.3%) |
| Has children | 51 (32.7%) | 16 (20.5%) | 28 (46.7%) | 53 (32.7%) | 104 (32.7%) |
| **Location** |  |  |  |  |  |
| San Francisco | 118 (75.6%) | 64 (82.1%) | 36 (60.0%) | 114 (70.4%) | 232 (73.0%) |
| Outside San Francisco | 38 (24.4%) | 14 (17.9%) | 24 (40.0%) | 48 (29.6%) | 86 (27.0%) |
| **Household Size** |  |  |  |  |  |
| 2 persons | 13 (8.3%) | 9 (11.5%) | 4 (6.7%) | 20 (12.3%) | 33 (10.4%) |
| 3-4 persons | 64 (41.0%) | 28 (35.9%) | 20 (33.3%) | 56 (34.6%) | 120 (37.7%) |
| 5+ persons | 79 (50.6%) | 41 (52.6%) | 36 (60.0%) | 86 (53.1%) | 165 (51.9%) |
| **Household Density\*** |  |  |  |  |  |
| Mean (SD) | 1.86 (0.858) | 1.92 (0.911) | 2.28 (1.21) | 2.04 (1.04) | 1.95 (0.956) |
| Median [Min, Max] | 1.88 [0.250, 7.00] | 1.63 [0.444, 5.00] | 2.00 [0.857, 6.00] | 1.71 [0.444, 6.00] | 1.75 [0.250, 7.00] |

<sup>+</sup> 22 households with S gene only sequence available.
\*Household density missing for 17 households.

**Table 2:** Secondary household attack rates for West Coast variants, combined and disaggregated by B.1.427 and B.1.429. Relative risks estimated based on Poisson regression using generalized estimating equations and cluster-robust standard errors. Adjustment variables included age

|  |  |  | Unadjusted |  | Adjusted |  |
| --- | --- | --- | --- | --- | --- | --- |
|  | **Positives**<br>**Among Tested**<br>**Contacts (%)** | **Mean**<br>**Household**<br>**Attack Rate**<br>**(95% CI)** | **RR (95%CI)** | **p-value** | **aRR** | **p-value** |
| **Class** |  |  |  |  |  |  |
| **Non-West**<br>**Coast** | 122/415<br>(29.4%) | 25.6% (20.3-31) | - | - | - | - |
| **West Coast** | 161/451<br>(35.7%) | 36.1% (30.3-42.2) | 1.29 (1.01-1.64) | 0.044 | 1.25 (0.98-1.60) | 0.068 |
| **Lineage** |  |  |  |  |  |  |
| **B.1.427** | 68/206 (33%) | 31.7% (23.6-40.1) | 1.18 (0.87-1.59) | 0.3 | 1.22 (0.90-1.64) | 0.2 |
| **B.1.429** | 77/186<br>(41.4%) | 42.2% (32.7-52.1) | 1.48 (1.11-1.98) | 0.008 | 1.39 (1.03-1.87) | 0.030 |

**Figure 1.**
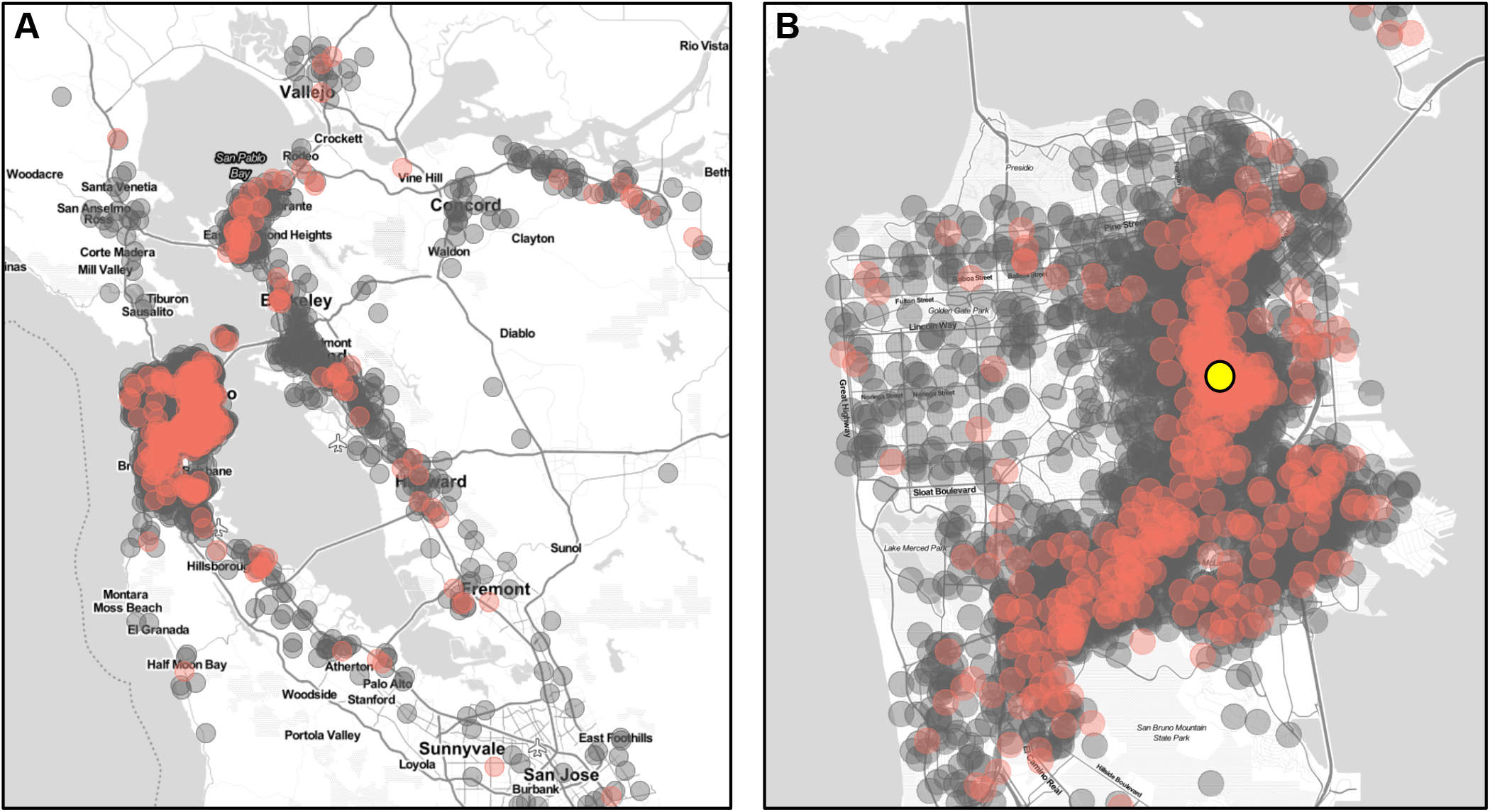
Testing catchment area. The location of the 24th & Mission testing site is denoted by the yellow symbol. Negative tests are in grey, and positive tests are shown in red. Household locations shown have a random offset of up to 750 meters. The testing catchment area is concentrated within San Francisco County (A), but substantial numbers of individuals reside in the surrounding 8 Bay Area Counties (B). Map tiles by Stamen Design and data by OpenStreetMap.

Similar to our previous observations in San Francisco[17], full length sequences were distributed among the major clades (Supplementary Figure 1). Notably, mutations at spike position 501 were not observed, and thus no instances of the B.1.1.7 strain or any other strain bearing the N501Y mutation were detected in any sample during this period. A single individual was found to have been infected with the P.2 strain, which carries the spike E484K mutation and was described in Brazil from a re-infection case[4]. This mutation has been associated with decreased neutralization in numerous laboratory experiments[1,3].

We observed SARS-CoV-2 genome sequences that belonged to PANGO lineages B.1.427 and B.1.429, both of which share a trio of recent mutations in the spike protein (S13I, W152C, and L452R) (Figure 2). These lineages are separated by differing mutations ORF1a and ORF1b, including ORF1b:P976L and ORF1a:I4205V, respectively. Sequencing of 191 viral genomes from November 22 – December 2, 2020 revealed that sequences carrying this trio of mutations represented only 15.7% of the total. A trend of increasing frequency was observed on a daily basis during the January testing period (Figure 2A), and the frequency of these lineages were observed to have increased to 54.9% of the total, representing an increase of more than 3-fold in approximately 1.5 months (Figure 2B,C). As noted, this increase in frequency is consistent with an expansion of viruses more broadly in California carrying these same mutations[18].

**Figure 2.**
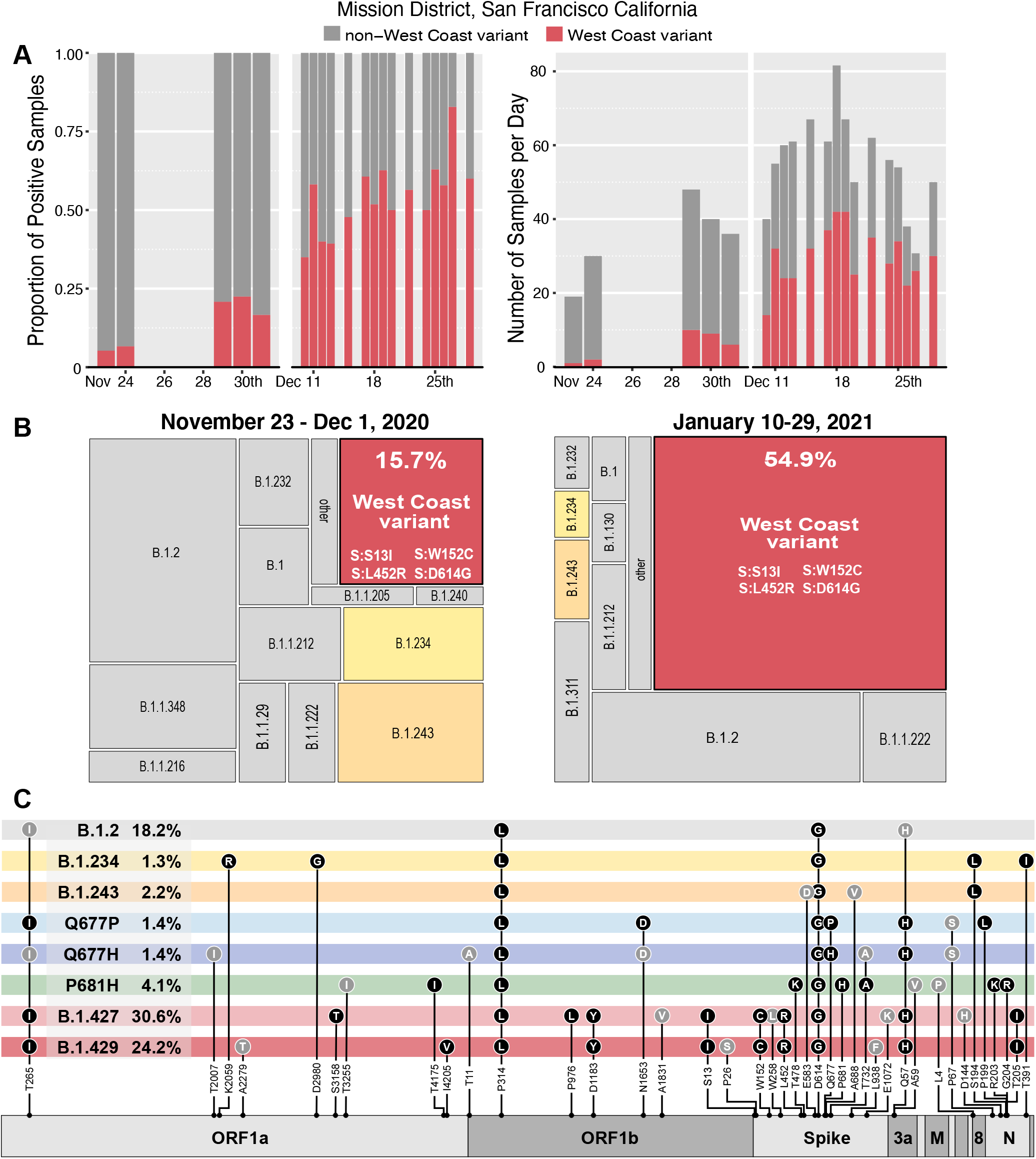
Variants observed at 24th & Mission. (A) The proportion of daily cases belonging to West Coast and non-West Coast variants, and (B) the total number of samples per day. (C,D) Area maps [25] showing the relative proportion of PANGO lineages acquired from full length genomes from the November (N=191) and January (N=671) time periods respectively. (E) Genome maps for variants detected in this study. Dominant mutations (filled black circles), and non-synonymous mutations detected at lower frequency in combination with existing lineages (filled grey circles) are shown in grey.

Additional non-synonymous mutations were observed throughout the genome, including 100 unique non-synonymous mutations in the spike gene, of which several are within functionally-significant regions of the protein (Figure 2C, Supplementary Table 3). Eleven unique mutations were observed in the receptor binding domain, most of which have yet to be investigated for possible effects on ACE2 binding, transmissibility, or antibody neutralization. Additionally, seven unique mutations were found adjacent to the polybasic furin cleavage site at the S1/S2 junction, which is reported to have a potential role in determination of virulence and host cell tropism[19–22]. Moderately prevalent mutations were observed at spike position 681 (P681H, n=29 and P681R, n=1), which is within the furin recognition site, and at spike position 677, where two different amino acid substitutions were observed in this cohort (Q677H, n=21 and Q677P, n=10). Multiple mutations at both of these sites have been observed across a wide geographical range with previous reports noting the likelihood of positive selection accounting for convergent evolution[6].

### Disease Severity

The SARS-CoV-2 RT-PCR cycle thresholds (Ct) for nasal swab samples from which whole genomes corresponding to the West Coast variant were recovered were compared to parallel non-West Coast variant samples. Mean Ct values did not differ significantly between persons infected with West Coast (mean Ct 23.62; IQR 6.6) versus non-West Coast (mean Ct 23.67; IQR 8.0) strains (95% CI: −0.63-0.83, p-value = 0.79) (Supplementary Figure 2, Supplementary Table 2). The proportion of individuals with symptoms was similar among persons infected with West Coast (272/447, 60.9%) versus non-West Coast (248/387, 64.1%) strains. Among 347 sequenced cases with longitudinal follow up by the Community Wellness Team, 4 (1.2%) were hospitalized (3/184, and 1/175, for West Coast and non-West Coast, respectively).

### Household Secondary Attack Rate

A total of 327 households met inclusion criteria for evaluation of secondary attack rate; of these, 9 households had individuals with mixed strains, and thus were excluded from analyses. Among the remaining 318 households, characteristics including race/ethnicity, ages of other household members, household size, density, and location were similar, regardless of whether the members were positive for West Coast or non-West Coast variants. (Table 1, Supplementary Table 4).

The 318 index cases had a total of 1,241 non-index household members; of these, 866 (69.8%) had a BinaxNOW test result available (451/655 [68.9%] for West Coast variant households; 415/586 [70.8%] of non-West Coast variant households). A total of 35.7% (161/451) of household contacts exposed to the West Coast variant tested BinaxNOW positive (33.0%, 68/206 for B1.427; 41.4%, 77/186 for B.1.429), while 29.4% (122/415) of contacts exposed to non-West Coast variant tested positive (Table 2). Secondary cases were identified a median of 1 day after index cases (IQR 0-4).

Based on unadjusted Poisson regression with cluster-robust standard errors, household contacts exposed to the West Coast variant had an estimated 29% higher risk of secondary infection, compared to household contacts exposed to a non-West Coast variant (RR: 1.29, 95% CI: 1.01-1.64, p-value = 0.044). When exposure to West Coast variants was disaggregated by B.1.427 and B.1.429, corresponding risks of secondary infections relative to exposure to non-West Coast variants were 1.18 (95% CI: 0.87-1.59, p-value = 0.3) and 1.48 (95% CI: 1.11-1.98, p-value = 0.008), respectively. Dispersion ratios were greater than 0.9 in all regression analyses. Estimated relative risks of infection after household exposure to West Coast versus non-West Coast variants were similar after adjustment for household and individual-level characteristics of secondary contacts (aRR: 1.25, 95% CI: 0.98-1.60, p-value: 0.068 for West Coast vs non-West Coast variants; aRR: 1.22, 95%CI: 0.90-1.64, p-value = 0.2 and aRR: 1.39, 95%CI: 1.03-1.87, p-value = 0.030 for B.1.427 and B.1.429, respectively.) Relative attack rates were generally similar when stratified by household characteristics and by the characteristics of secondary contacts (Table 3); secondary attack rates among children aged <12 years were 51.9% (41/79) and 39.7% (31/78) when exposed to West Coast and non-West Coast strains, respectively. Sensitivity analyses in which parametric assumptions were relaxed using TMLE and Super Learning yielded similar estimates (Supplementary Table 5).

**Table 3:** Secondary attack rate disaggregated by covariates; mean household secondary attack rate only reported disaggregated by household level characteristics.

|  | Non-West Coast Strain |  | West Coast Strain |  |
| --- | --- | --- | --- | --- |
|  | Positives Among Tested Contacts (%) | Mean Household Attack Rate (95% CI) | Positives Among Tested Contacts (%) | Mean Household Attack Rate (95% CI) |
| **Location** |  |  |  |  |
| San Francisco | 88/321 (27.4%) | 22.9% (17.2-28.8) | 113/315 (35.9%) | 37.8% (30.6-45.1) |
| Outside of San Francisco | 34/94 (36.2%) | 34% (22.1-46.2) | 48/136 (35.3%) | 32.1% (22.2-42.1) |
| **Age Group** |  |  |  |  |
| Age ≤ 12 | 31/78 (39.7%) | - | 41/79 (51.9%) | - |
| Age > 12 | 91/337 (27%) | - | 120/372 (32.3%) | - |
| **Race/Ethnicity** |  |  |  |  |
| Latinx/Hispanic | 107/372 (28.8%) | - | 136/378 (36%) | - |
| Not Latinx/Hispanic | 15/43 (34.9%) | - | 25/73 (34.2%) | - |
| **Household Size** |  |  |  |  |
| 2 persons | 1/13 (7.7%) | 7.7% (0-23.1) | 12/20 (60%) | 60% (40-80) |
| 3-4 persons | 30/116 (25.9%) | 26% (17.4-35.4) | 35/101 (34.7%) | 33.9% (24.1-44.3) |
| 5+ persons | 91/286 (31.8%) | 28.1% (21.1-35.4) | 114/330 (34.5%) | 32% (25.1-39.2) |
| **Household Density** |  |  |  |  |
| Bottom half | 43/160 (26.9%) | 23.4% (15.4-31.6) | 52/175 (29.7%) | 33.4% (24.3-42.9) |
| Top half | 76/242 (31.4%) | 27.7% (20.5-35.3) | 101/262 (38.5%) | 35.7% (27.9-43.6) |

### Estimation of Reproductive Number

Using Bayesian phylogenetic analysis, we estimated the reproductive number to be 1.27 (95% CI: 1.10-1.46) for B.1.427 and 1.18 (95% CI: 1.05-1.32) for B.1.429 during the second half of 2020. These values were slightly higher than two other lineages spreading in California during the same time period: 1.12 (95% CI: 1.10-1.14) for B.1.232, and 1.02 (95% CI: 0.98-1.05) for B.1.243. As the reproductive numbers are very similar and were calculated from the median SkyGrid estimates, we cannot conclude any statistically significant differences between the lineages.

## DISCUSSION

We monitored SARS-CoV-2 viral variants by genomic sequencing and integration of metadata from households at a community based “test-and-respond” program. We found that the West Coast variants (PANGO lineages B.1.427 and B.1.429) increased in prevalence relative to wild type from November to January in the San Francisco Bay Area among persons tested in the same community-based location. These data extend and confirm prior observations from convenience, outbreak, and clinical samples reporting apparent increases in relative prevalence of the West Coast variants[18].

Household secondary attack rates of the West Coast variants were modestly higher than for non-West Coast variants, suggesting the potential for increased transmissibility. The West Coast variants compromise two closely related lineages (B.1.427 and B.1.429) that share identical sets of mutations in the spike protein, but differ by additional synonymous and non-synonymous mutations in other genes, including ORF1a and ORF1b. While the frequency of both lineages increased in this study and in California more widely[18], and the estimated increase in risk of secondary household infection relative to non-West Coast variants was fairly consistent across lineages, the point estimate was somewhat higher for B.1.429. Although moderate compared to increased transmissibility of some other previously identified variants, even small increases in transmissibility could contribute to a substantial increase in cases, particularly in the context of reproductive numbers just below one. While this finding may be due to chance, future work, should continue to monitor individual lineages.

The household attack rate observed here was higher than that reported in a recent global meta analysis[23], even for the non-West Coast variants. It was similar to, or lower than attack rates reported in other US settings. Prior US reports, however, were based on substantially smaller sample sizes.

Our findings that the West Coast variants increased in relative prevalence and had higher household secondary attack rates potentially suggest higher transmissibility. However, the West Coast variant has been detected in multiple locations, and has been detected since May 2020 in California without relative expansion until the peak associated with the holiday season of November-January. Using Bayesian phylogenetic analysis, the estimated reproductive number for both West Coast lineages was found to be modestly higher than other circulating lineages. However, further studies are required to evaluate the contribution of specific mutations.

We found no significant differences in viral load (using Ct) between West Coast and non-West Coast variants (Supplementary Figure 2), and recorded hospitalizations (n=5/388) remained rare, despite the West Coast variant representing 54.9% of positive cases. This highlights the importance of studying walk-up populations, whether they are symptomatic or asymptomatic, as hospitalized populations often are confounded by co-morbidities and subject to selection bias.

At the time of this sampling, no instances of B.1.1.7, or independent N501Y mutations were detected in our sample population of 830, despite sporadic observations elsewhere in CA (approximately 3% [69/2423] of genomes reported in California during the January study period; accessed from GISAID Feb 24, 2021). This suggests that introductions of B.1.1.7 to the sample population may not yet have occurred or have been rare[24]. A single case of the P.2 variant, which carries the E484K mutation[1], was detected in this study. Surprisingly, this case did not have a travel history, highlighting the risk of cryptic transmission.

In addition to the mutations associated with spike L452R in the West Coast variants, we observed, at lower frequencies, other mutations of interest, including those in spike at positions 677, and 681, both of which have been reported previously on their own, or in the case of 681, in the context of other mutations such as N501Y[6].

This study has several limitations. First, testing was conducted at a walk-up testing site, and thus these are inherently convenience samples; however, this would not be expected to impose a differential selection bias for those with or without any particular variant. Second, clear classification of the index case was not always possible, particularly when multiple adults from a household tested positive on the same date; further, secondary household attack rate calculations do not account for potential external sources of infection other than the index case. However, the relative risk of secondary infection from household exposure to West Coast versus non-West Coast variants was similar among children, a group less likely to have been misclassified as non-index or to be exposed to external infection. Third, household testing coverage was incomplete; this might contribute to an over (or under) estimate of secondary attack rate, and while we again have no reason to suspect differential ascertainment by strain, this could bias estimates of relative risk.

The occurrence of variants in SARS-CoV-2 was always expected; however, it is often difficult to understand the importance of any given single or co-occurring mutations. While further epidemiological and laboratory experiments will be required to fully understand the community impact and mechanistic underpinnings of each variant, it is clear that enhanced genomic surveillance, paired with community engagement, testing, and response capacity is an important tool in the arsenal against this pandemic.

## Data Availability

Genomic sequences have been deposited in GISAID.

https://www.gisaid.org

## ACKNOWLEDGMENTS

We would like to thank the hundreds of academic labs and public health institutions that have been sequencing and publicly depositing genomic sequences, analysis tools, and epidemiological data throughout the pandemic. We thank Bevan Dufty and the BART team, Jeff Tumlin and the San Francisco MUNI, Supervisor Hillary Ronen, Mayor London Breed, Dr. Grant Colfax and the Department of Public Health, Salu Ribeiro and Bay Area Phlebotomy and Laboratory services, PrimaryBio COVID testing platform, and our community ambassadors and volunteers. We would like to thank the Chan Zuckerberg Initiative, Greg and Lisa Wendt, Anne-Marie and Wylie Peterson, Gwendolyn Holcombe, Kevin and Julia Hartz, and Carl Kawaja for their critical support and input. We also thank Jack Kamm, Peter Kim, Don Ganem, Sandy Schmidt, Cori Bargmann, Norma Neff, and Christopher Hoover for technical assistance and discussion.

## FIGURE LEGENDS

**Supplementary Figure S1:**
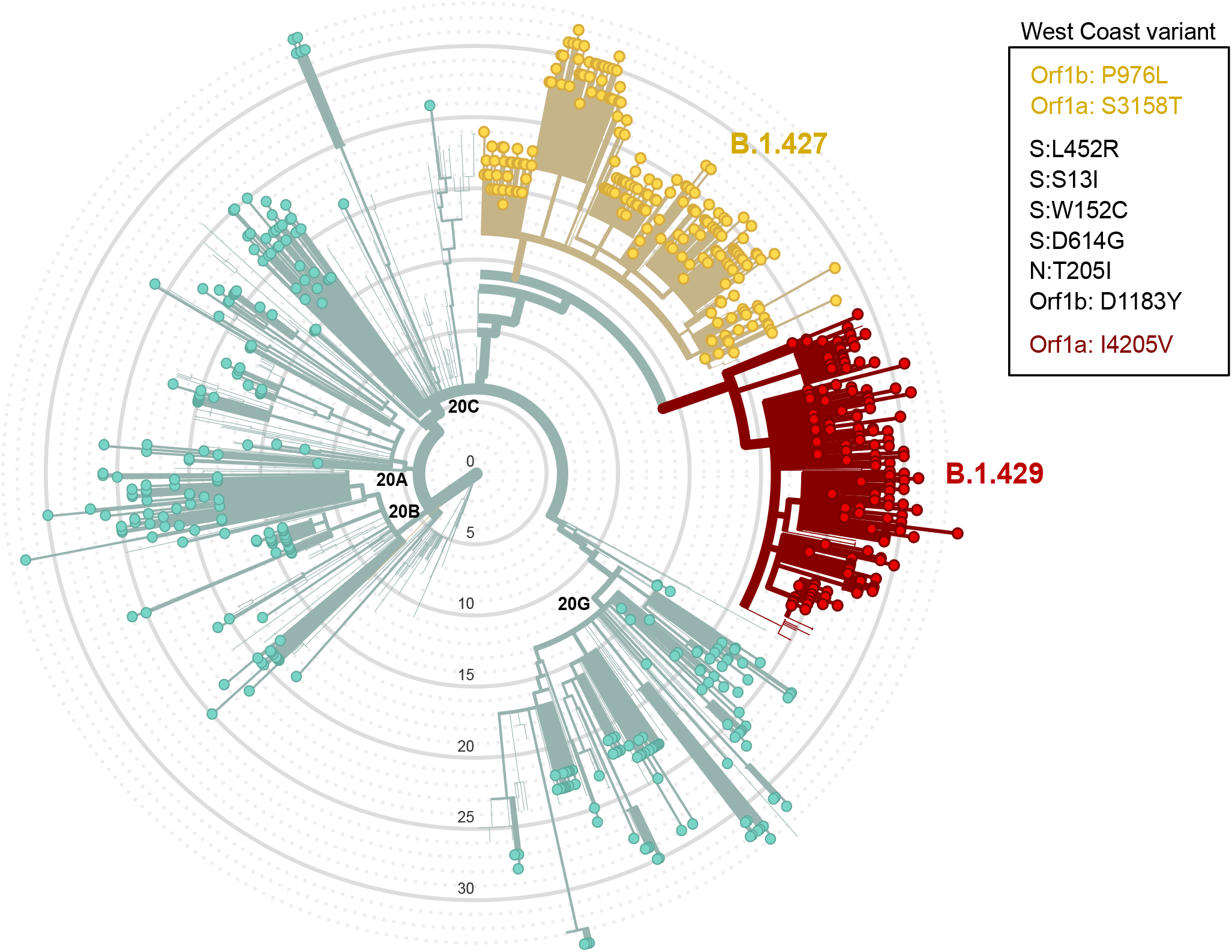
Nextclade Phylogeny. Radial tree phylogeny as visualized via Nextclade [26] using full length genomes (n=614) from the Jan 10th-29th sampling period and the San Francisco public reference tree (downloaded Feb 17, 2021). For the purpose of visualization, six genomes were removed due to excessive private mutations. Colored nodes correspond to specific mutations as indicated in the figure legend.

**Supplementary Figure S2:**
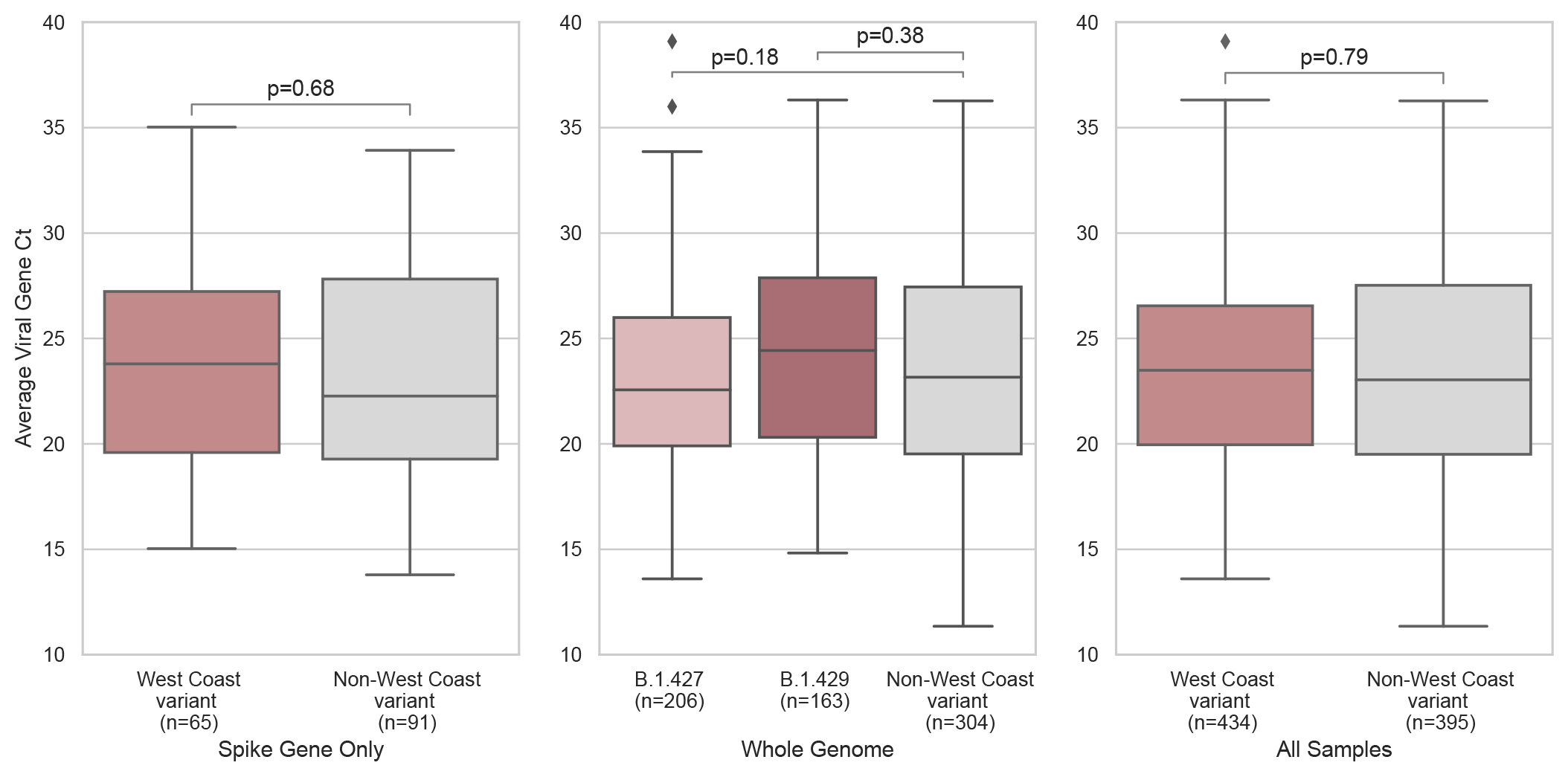
Viral load by Ct value. No significant differences in mean viral load as measured by RT-qPCR Cts was measured between West Coast and non-West Coast variants in (A) the subset of samples that had only spike gene sequenced (95% CI: −1.97-1.29, p-value = 0.68), (B) the subset of samples that had undergone whole genome sequencing thus allowing for determination of West Coast variant PANGO lineage of either B.1.427 (95% CI: −1.84-0.25, p-value = 0.18) or B.1.429 (95% CI: −0.49-1.76, p-value = 0.38), (C) all samples sequenced in this study (95% CI: −0.63-0.83, p-value = 0.79).

**Supplementary Table S1:** Characteristics of all persons (N=8,822; 10.0% with positive result) tested at the community-based testing site between January 10-29.

|  | Negative (N=7937) | Positive (N=885) | Total (N=8822) |
| --- | --- | --- | --- |
| **Gender** |  |  |  |
| Female | 3595 (90.2%) | 391 (9.8%) | 3986 (100%) |
| Male | 4190 (89.7%) | 481 (10.3%) | 4671 (100%) |
| Other | 152 (92.1%) | 13 (7.9%) | 165 (100%) |
| **Age Group** |  |  |  |
| Age ≤ 12 | 488 (83.1%) | 99 (16.9%) | 587 (100%) |
| Age 13-35 | 3032 (88.8%) | 382 (11.2%) | 3414 (100%) |
| Age 36-64 | 3844 (91.5%) | 355 (8.5%) | 4199 (100%) |
| Age ≥ 65 | 573 (92.1%) | 49 (7.9%) | 622 (100%) |
| **Race/Ethnicity** |  |  |  |
| Hispanic/Latinx | 5700 (88.5%) | 744 (11.5%) | 6444 (100%) |
| Asian | 610 (93.7%) | 41 (6.3%) | 651 (100%) |
| White/Caucasian | 818 (96.3%) | 31 (3.7%) | 849 (100%) |
| Black or African American | 228 (96.6%) | 8 (3.4%) | 236 (100%) |
| Other | 581 (90.5%) | 61 (9.5%) | 642 (100%) |
| **Occupation\*** |  |  |  |
| Food & Beverage | 987 (90.5%) | 104 (9.5%) | 1091 (100%) |
| Tradesperson | 423 (87%) | 63 (13%) | 486 (100%) |
| Day laborer | 227 (89.4%) | 27 (10.6%) | 254 (100%) |
| Healthcare | 277 (93.9%) | 18 (6.1%) | 295 (100%) |
| Student | 780 (84.4%) | 144 (15.6%) | 924 (100%) |
| Other | 2557 (92.4%) | 211 (7.6%) | 2768 (100%) |
| No employment | 1126 (88.8%) | 142 (11.2%) | 1268 (100%) |
| **Day of Test Symptoms** |  |  |  |
| Asymptomatic | 6089 (94.6%) | 347 (5.4%) | 6436 (100%) |
| Symptomatic | 1848 (77.5%) | 538 (22.5%) | 2386 (100%) |

\* Occupation information missing from 1,736 persons.

**Supplementary Table S2:** Sample sequencing summary and viral cycle threshold characteristics.

|  |  | Spike Gene Only | Whole Genome |  | Total Count (Jan 10-Jan 30) |
| --- | --- | --- | --- | --- | --- |
| **Total Samples Run** |  | 165 | 674 |  | 839 |
| **Total High Quality (>92% coverage)** |  | 96 | 620 |  | 716 |
| **Average Reads/Sample** |  | 96,237 | 2,437,080 |  | - |
| **Average % Gene/Genome Covered** |  | 83.35 | 97.09 |  | 94.39 |
| **Mean Viral Ct** |  | 23.78 | 23.63 |  | 23.65 |
| Mean Viral Ct<br>West Coast variant | B.1.427 | 24.00 | 23.60 | 22.96 | 23.62 |
|  | B.1.429 |  |  | 24.40 |  |
| Mean Viral Ct<br>Non-West Coast variant |  | 23.63 | 23.76 |  | 23.67 |
| **Median Viral Ct** |  | 23.65 | 23.26 |  | 23.26 |
| Median Viral Ct<br>West Coast variant | B.1.427 | 24.00 | 23.42 | 22.56 | 23.49 |
|  | B.1.429 |  |  | 24.42 |  |
| Median Viral Ct<br>Non-West Coast variant |  | 22.83 | 23.16 |  | 23.04 |
| **Viral Ct Interquartile Range (IQR)** |  | 8.29 | 7.16 |  | 7.42 |
| Viral Ct IQR<br>West Coast variant | B.1.427 | 7.39 | 6.40 | 6.09 | 6.59 |
|  | B.1.429 |  |  | 7.57 |  |
| Viral Ct IQR Non-West Coast variant |  | 8.88 | 7.93 |  | 8.01 |

**Supplementary Table S3:**
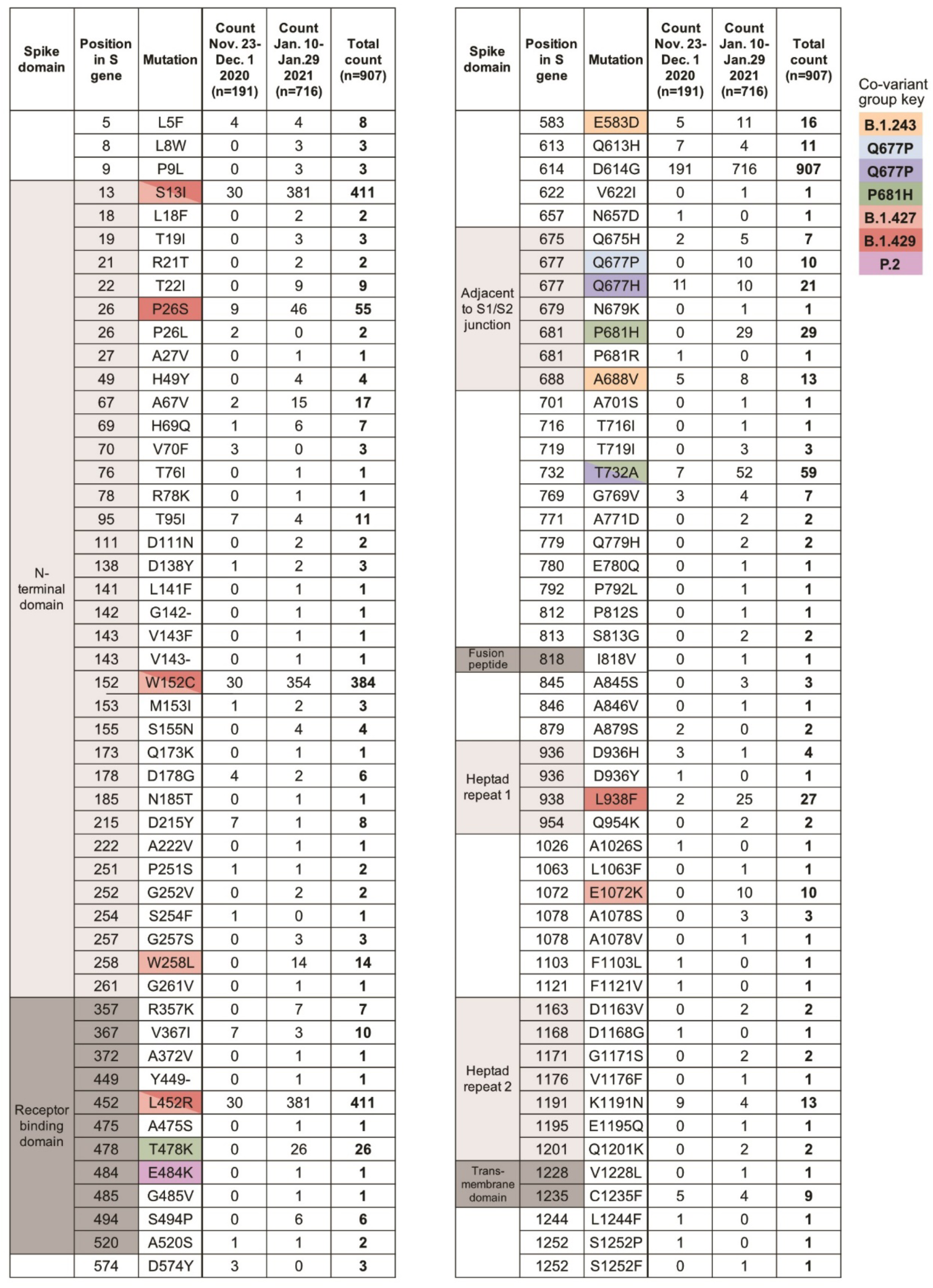
Amino acid substitutions observed in the spike gene and count of sequences per mutation in each study.

**Supplementary Table S4:** Individual characteristics of all persons tested (both positive and negative, and including index case) living in one of the 318 households meeting inclusion criteria for household secondary attack rate analyses, stratified by strain classification of the household.

|  | Non-West coast<br>(N=571) | West coast |  |  | Total<br>(N=1184) |
| --- | --- | --- | --- | --- | --- |
|  |  | B.1.427<br>(N=284) | B.1.429<br>(N=246) | All West coast<br>(N=613) |  |
| **Sex** |  |  |  |  |  |
| Female | 275 (48.2%) | 123 (43.3%) | 120 (48.8%) | 286 (46.7%) | 561 (47.4%) |
| Male | 288 (50.4%) | 156 (54.9%) | 124 (50.4%) | 320 (52.2%) | 608 (51.4%) |
| Other | 8 (1.4%) | 5 (1.8%) | 2 (0.8%) | 7 (1.1%) | 15 (1.3%) |
| **Age Group** |  |  |  |  |  |
| Age <= 12 | 78 (13.7%) | 24 (8.5%) | 42 (17.1%) | 79 (12.9%) | 157 (13.3%) |
| Age 13-35 | 253 (44.3%) | 117 (41.2%) | 105 (42.7%) | 257 (41.9%) | 510 (43.1%) |
| Age 36-64 | 216 (37.8%) | 126 (44.4%) | 90 (36.6%) | 245 (40.0%) | 461 (38.9%) |
| Age >= 65 | 24 (4.2%) | 17 (6.0%) | 9 (3.7%) | 32 (5.2%) | 56 (4.7%) |
| **Race/Ethnicity** |  |  |  |  |  |
| Hispanic/Latinx | 511 (89.5%) | 233 (82.0%) | 217 (88.2%) | 518 (84.5%) | 1029<br>(86.9%) |
| Asian | 17 (3.0%) | 10 (3.5%) | 12 (4.9%) | 28 (4.6%) | 45 (3.8%) |
| White/Caucasian | 16 (2.8%) | 13 (4.6%) | 3 (1.2%) | 18 (2.9%) | 34 (2.9%) |
| Black or African American | 4 (0.7%) | 7 (2.5%) | 1 (0.4%) | 12 (2.0%) | 16 (1.4%) |
| Other | 23 (4.0%) | 21 (7.4%) | 13 (5.3%) | 37 (6.0%) | 60 (5.1%) |
| **Occupation\*** |  |  |  |  |  |
| Food & Beverage | 68 (11.9%) | 48 (16.9%) | 23 (9.3%) | 80 (13.1%) | 148 (12.5%) |
| Tradesperson | 35 (6.1%) | 16 (5.6%) | 4 (1.6%) | 24 (3.9%) | 59 (5.0%) |
| Day laborer | 19 (3.3%) | 8 (2.8%) | 9 (3.7%) | 18 (2.9%) | 37 (3.1%) |
| Healthcare | 13 (2.3%) | 7 (2.5%) | 2 (0.8%) | 11 (1.8%) | 24 (2.0%) |
| Student | 110 (19.3%) | 34 (12.0%) | 62 (25.2%) | 110 (17.9%) | 220 (18.6%) |
| Other | 133 (23.3%) | 70 (24.6%) | 62 (25.2%) | 148 (24.1%) | 281 (23.7%) |
| No employment | 86 (15.1%) | 55 (19.4%) | 43 (17.5%) | 115 (18.8%) | 201 (17.0%) |

| **Day of Test**<br>**Symptoms** |  |  |  |  |  |
| --- | --- | --- | --- | --- | --- |
| Asymptomatic | 320 (56.0%) | 166 (58.5%) | 131 (53.3%) | 348 (56.8%) | 668 (56.4%) |
| Symptomatic | 251 (44.0%) | 118 (41.5%) | 115 (46.7%) | 265 (43.2%) | 516 (43.6%) |

\* Occupation information missing for 214 persons.

**Supplementary Table S5:** Adjusted attack rates from sensitivity analysis using Targeted Maximum Likelihood and Super Learning.

|  | Adjusted |  |
| --- | --- | --- |
|  | aRR (95% CI) | p-value |
| **Class** |  |  |
| **Non-West Coast** | - | - |
| **West Coast** | 1.17 (0.93-1.49) | 0.188 |
| **Lineage** |  |  |
| **B.1.427** | 1.17 (0.87-1.56) | 0.294 |
| **B.1.429** | 1.34 (1.01-1.78) | 0.045 |

## SUPPLEMENTARY METHODS

### SARS-CoV-2 genomic sequence recovery

Swab samples from individuals testing positive by BinaxNOW were placed in DNA/RNA shield and processed as previously described [1]. Extracted total nucleic acid was diluted based on average SARS-CoV-2 N and E gene cycle threshold (Ct) values; samples with a Ct range 12-15 were diluted 1:100, 15-18 1:10 and >18 no dilution. For high throughput scaling, library preparation reaction volumes and dilutions were miniaturized utilizing acoustic liquid handling. Library preparation followed either the modified versions of the Primal-Seq Nextera XT version 2.0 protocol [2,3], or the modified version of SARS-CoV-2 Tailed Amplicon Illumina Sequencing V.2 [4], both using the ARTIC Network V3 primers [5]. A subset of initial samples were library prepared using the Tailed Amplicon Sequencing V.2 with only primer pairs 71-84 of the ARTIC V3 primers to tile all of the S gene. Final libraries were sequenced by paired-end 2 x 150bp sequencing on an Illumina NovaSeq platform, or for the S-gene-only set, 2 x 300bp on an Illumina MiSeq.

### SARS-CoV-2 consensus genome generation

Raw .fastq files were imported into IDseq and consensus genomes were generated automatically using the embedded SARS-CoV-2 pipeline [6]. Specifically, minimap2 was used to align raw reads to the reference genome MN908947.2 [7], then the consensus sequence was generated using samtools [8], mpileup and ivar [9]. The IDseq consensus genome pipeline is implemented in WDL [10]. Viral genomes with at least 92% (27,500nt) recovery were uploaded to GISAID [11], a worldwide repository for SARS-CoV-2 genomes and Genbank [12]. Phylogenetic analysis was conducted and results were visualized in Nextclade (https://clades.nextstrain.org) [13].

### Bayesian Phylogenetic Analysis

To compare the viral diversity of the two variants of interest, B.1.427 and B.1.429, we identified two other SARS-CoV-2 lineages, B.1.232 and B.1.243 that were prevalent in the state of California from July 2020 onwards. Both of these lineages contained more sequences on GISAID from California than any other location, based on the PANGO lineage assignment [14] on GISAID. For each of the 4 lineages spreading in California, we randomly subsampled all available genomes from GISAID and this study to 500 or fewer genomes. For subsampled genomes, we aligned them against the reference genome (Genbank accession: MN996528.1) using MAFFT v7.471 [15] with default settings. Each multi-sequence alignment was used to build a separate maximum likelihood tree in IQ-TREE v.1.6.12 [16] with default options. The trees were rooted at the reference genome. The maximum likelihood tree was used to visually identify outlier sequences which could have been misclassified as that PANGO lineage. The resulting number of genomes included in the downstream analysis were: B.1.232: 368; B.1.243: 500; B.1.427: 495; B.1.429: 443. The multi-sequence alignment of the coding region for each lineage was analyzed in BEAST v.1.10.4 [17] with unlinked molecular clocks between the S gene and other genes, uncorrelated relaxed molecular clock (lognormal distribution), GTR substitution model with 4 rate categories (selected by the BIC value in ModelTest [18]), and the Bayesian Skygrid population model [19]. Default prior values and operator values were used. The MCMC chains were 100M in length, sampled every 50,000, and the first 50% of samples discarded as burnin. We ran 2 replicate MCMC chains for each analysis, and used all samples to summarize the results.

We fit an exponential model to the median SkyGrid estimate between 2020-07-01 and 2021-01-01 to calculate the growth rate per day *r*, and calculated the reproductive number *R* using the formula [20] R = (1+r/b)^a^, where a=1.39 is the shape parameter and b=0.14 is the scale parameter of a gamma generation time distribution with a mean of 5 days a standard deviation of 1.9 days [21]. The 95% confidence interval (95% CI) around *R* was calculated based on the 95% CI around the *r* estimate. Samples used for the Bayesian Analysis are detailed in the supplementary file.

## REFERENCES

1. Liu Z, VanBlargan LA, Bloyet L-M, et al. Landscape analysis of escape variants identifies SARS-CoV-2 spike mutations that attenuate monoclonal and serum antibody neutralization. BioRxiv Prepr Serv Biol 2020;

2. Voloch CM, F R da S, Almeida LGP de, et al. Genomic characterization of a novel SARS-CoV-2 lineage from Rio de Janeiro, Brazil. medRxiv 2020; :2020.12.23.20248598.

3. Weisblum Y, Schmidt F, Zhang F, et al. Escape from neutralizing antibodies by SARS-CoV-2 spike protein variants. bioRxiv 2020; :2020.07.21.214759.

4. Ferrareze PAG, Franceschi VB, Mayer A de M, Caldana GD, Zimerman RA, Thompson CE. E484K as an innovative phylogenetic event for viral evolution: Genomic analysis of the E484K spike mutation in SARS-CoV-2 lineages from Brazil. bioRxiv 2021; :2021.01.27.426895.

5. Zeller M, Gangavarapu K, Anderson C, et al. Emergence of an early SARS-CoV-2 epidemic in the United States. medRxiv 2021; :2021.02.05.21251235.

6. Hodcroft EB, Domman DB, Oguntuyo K, et al. Emergence in late 2020 of multiple lineages of SARS-CoV-2 Spike protein variants affecting amino acid position 677. medRxiv 2021; :2021.02.12.21251658.

7. 12.20.20_CCSF_Press_Release.pdf. Available at: https://www.sfdph.org/dph/alerts/files/12.20.20_CCSF_Press_Release.pdf. Accessed 24 February 2021.

8. Pilarowski G, Marquez C, Rubio L, et al. Field performance and public health response using the BinaxNOW TM Rapid SARS-CoV-2 antigen detection assay during community-based testing. Clin Infect Dis Off Publ Infect Dis Soc Am 2020; Available at: https://www.ncbi.nlm.nih.gov/pmc/articles/PMC7799223/. Accessed 23 February 2021.

9. Pilarowski G, Lebel P, Sunshine S, et al. Performance Characteristics of a Rapid Severe Acute Respiratory Syndrome Coronavirus 2 Antigen Detection Assay at a Public Plaza Testing Site in San Francisco. J Infect Dis 2021; Available at: https://doi.org/10.1093/infdis/jiaa802. Accessed 1 March 2021.

10. Rubio LA, Peng J, Rojas S, et al. The COVID-19 Symptom to Isolation Cascade in a Latinx Community: A Call to Action. Open Forum Infect Dis 2021; 8:ofab023.

11. Kerkhoff AD, Sachdev D, Mizany S, et al. Evaluation of a novel community-based COVID-19 ‘Test-to-Care’ model for low-income populations. PloS One 2020; 15:e0239400.

12. artic-network/artic-ncov2019. Available at: https://github.com/artic-network/artic-ncov2019. Accessed 25 February 2021.

13. Kalantar KL, Carvalho T, de Bourcy CFA, et al. IDseq-An open source cloud-based pipeline and analysis service for metagenomic pathogen detection and monitoring. GigaScience 2020; 9.

14. Gelman A, Hill J. Data analysis using regression and multilevel/hierarchical models. Cambridge?; New York: Cambridge University Press, 2007.

15. Laan MJ van der, Rose S. Targeted Learning in Data Science: Causal Inference for Complex Longitudinal Studies. Springer International Publishing, 2018. Available at: https://www.springer.com/gp/book/9783319653037. Accessed 24 February 2021.

16. Elbe S, Buckland-Merrett G. Data, disease and diplomacy: GISAID’s innovative contribution to global health. Glob Chall Hoboken NJ 2017; 1:33–46.

17. Chamie G, Marquez C, Crawford E, et al. SARS-CoV-2 Community Transmission disproportionately affects Latinx population during Shelter-in-Place in San Francisco. Clin Infect Dis Off Publ Infect Dis Soc Am 2020; Available at: https://www.ncbi.nlm.nih.gov/pmc/articles/PMC7499499/. Accessed 24 February 2021.

18. Zhang W, Davis BD, Chen SS, Martinez JMS, Plummer JT, Vail E. Emergence of a novel SARS-CoV-2 strain in Southern California, USA. medRxiv 2021; :2021.01.18.21249786.

19. Hoffmann M, Kleine-Weber H, Pöhlmann S. A Multibasic Cleavage Site in the Spike Protein of SARS-CoV-2 Is Essential for Infection of Human Lung Cells. Mol Cell 2020; 78:779–784.e5.

20. Örd M, Faustova I, Loog M. The sequence at Spike S1/S2 site enables cleavage by furin and phospho-regulation in SARS-CoV2 but not in SARS-CoV1 or MERS-CoV. Sci Rep 2020; 10:16944.

21. Johnson BA, Xie X, Bailey AL, et al. Loss of furin cleavage site attenuates SARS-CoV-2 pathogenesis. Nature 2021; :1–7.

22. Jaimes JA, Millet JK, Whittaker GR. Proteolytic Cleavage of the SARS-CoV-2 Spike Protein and the Role of the Novel S1/S2 Site. iScience 2020; 23:101212.

23. Madewell ZJ, Yang Y, Longini IM, Halloran ME, Dean NE. Household Transmission of SARS-CoV-2: A Systematic Review and Meta-analysis. JAMA Netw Open 2020; 3:e2031756.

24. Volz E, Mishra S, Chand M, et al. Transmission of SARS-CoV-2 Lineage B.1.1.7 in England: Insights from linking epidemiological and genetic data. medRxiv 2021; :2020.12.30.20249034.

25. Laserson U. laserson/squarify. 2021. Available at: https://github.com/laserson/squarify. Accessed 26 February 2021.

26. Hadfield J, Megill C, Bell SM, et al. Nextstrain: real-time tracking of pathogen evolution. Bioinforma Oxf Engl 2018; 34:4121–4123.

## REFERENCES

1. Crawford ED, Acosta I, Ahyong V, et al. Rapid deployment of SARS-CoV-2 testing: The CLIAHUB. PLOS Pathog 2020; 16:e1008966.

2. Quick J, Grubaugh ND, Pullan ST, et al. Multiplex PCR method for MinION and Illumina sequencing of Zika and other virus genomes directly from clinical samples. Nat Protoc 2017; 12:1261–1276.

3. R&d DP. COVID-19 ARTIC v3 Illumina library construction and sequencing protocol. 2020; Available at: https://www.protocols.io/view/covid-19-artic-v3-illumina-library-construction-an-bibtkann. Accessed 23 February 2021.

4. Gohl DM, Garbe J, Grady P, et al. A rapid, cost-effective tailed amplicon method for sequencing SARS-CoV-2. BMC Genomics 2020; 21:863.

5. artic-network/artic-ncov2019. Available at: https://github.com/artic-network/artic-ncov2019. Accessed 25 February 2021.

6. Kalantar KL, Carvalho T, de Bourcy CFA, et al. IDseq-An open source cloud-based pipeline and analysis service for metagenomic pathogen detection and monitoring. GigaScience 2020; 9.

7. Li H. Minimap2: pairwise alignment for nucleotide sequences. Bioinforma Oxf Engl 2018; 34:3094–3100.

8. Li H, Handsaker B, Wysoker A, et al. The Sequence Alignment/Map format and SAMtools. Bioinforma Oxf Engl 2009; 25:2078–2079.

9. Grubaugh ND, Gangavarapu K, Quick J, et al. An amplicon-based sequencing framework for accurately measuring intrahost virus diversity using PrimalSeq and iVar. Genome Biol 2019; 20:8.

10. chanzuckerberg/idseq-workflows. Chan Zuckerberg Initiative, 2021. Available at: https://github.com/chanzuckerberg/idseq-workflows. Accessed 25 February 2021.

11. Elbe S, Buckland-Merrett G. Data, disease and diplomacy: GISAID’s innovative contribution to global health. Glob Chall Hoboken NJ 2017; 1:33–46.

12. Clark K, Karsch-Mizrachi I, Lipman DJ, Ostell J, Sayers EW. GenBank. Nucleic Acids Res 2016; 44:D67–72.

13. Hadfield J, Megill C, Bell SM, et al. Nextstrain: real-time tracking of pathogen evolution. Bioinforma Oxf Engl 2018; 34:4121–4123.

14. Rambaut A, Holmes EC, O’ TooleÁ, et al. A dynamic nomenclature proposal for SARS-CoV-2 lineages to assist genomic epidemiology. Nat Microbiol 2020; 5:1403–1407.

15. Katoh K, Standley DM. MAFFT multiple sequence alignment software version 7: improvements in performance and usability. Mol Biol Evol 2013; 30:772–780.

16. Nguyen L-T, Schmidt HA, von Haeseler A, Minh BQ. IQ-TREE: a fast and effective stochastic algorithm for estimating maximum-likelihood phylogenies. Mol Biol Evol 2015; 32:268–274.

17. Suchard MA, Lemey P, Baele G, Ayres DL, Drummond AJ, Rambaut A. Bayesian phylogenetic and phylodynamic data integration using BEAST 1.10. Virus Evol 2018; 4:vey016.

18. Posada D, Crandall KA. MODELTEST: testing the model of DNA substitution. Bioinforma Oxf Engl 1998; 14:817–818.

19. Gill MS, Lemey P, Faria NR, Rambaut A, Shapiro B, Suchard MA. Improving Bayesian population dynamics inference: a coalescent-based model for multiple loci. Mol Biol Evol 2013; 30:713–724.

20. Fraser C, Donnelly CA, Cauchemez S, et al. Pandemic potential of a strain of influenza A (H1N1): early findings. Science 2009; 324:1557–1561.

21. Ferretti L, Wymant C, Kendall M, et al. Quantifying SARS-CoV-2 transmission suggests epidemic control with digital contact tracing. Science 2020; 368.

